# Predicted effects of summer holidays and seasonality on the SARS-Cov-2 epidemic in France

**DOI:** 10.1101/2020.07.06.20147660

**Authors:** Louis Duchemin, Mathilde Paris, Bastien Boussau

**Affiliations:** Université de Lyon, Université Lyon 1, CNRS, Laboratoire de Biométrie et Biologie Evolutive UMR 5558, F-69622 Villeurbanne, France; IGFL, Institut de Génomique Fonctionnelle de Lyon, Université de Lyon, Ecole Normale Supérieure de Lyon, CNRS, Université Claude Bernard Lyon 1, UMR 5242, 69364 Cedex 07, Lyon, France

## Abstract

The SARS-CoV-2 epidemic in France has had a large death toll. It has not affected all regions similarly, since the death rate can vary several folds between regions where the epidemic has remained at a low level and regions where it got an early burst. The epidemic has been slowed down by a lockdown that lasted for almost eight weeks, and individuals can now move between metropolitan French regions without restriction. In this report we investigate the effect on the epidemic of summer holidays, during which millions of individuals will move between French regions. Additionally, we evaluate the effect of strong or weak seasonality and of several values for the reproduction number on the epidemic, in particular on the timing, the height and the spread of a second wave. To do so, we extend a SEIR model to simulate the effect of summer migrations between regions on the number and distribution of new infections. We find that the model predicts little effect of summer migrations on the epidemic, because the number of migrating infectious individuals are low as a consequence of the lockdown. However, all the reproduction numbers above 1.0 and the seasonality parameters we tried result in a second epidemic wave, with a peak date that can vary between October 2020 and April 2021. If the sanitary measures currently in place manage to keep the reproduction number below 1.0, the second wave will be avoided. If they keep the reproduction number at a low value, for instance at 1.1 as in one of our simulations, the second wave is flattened and could be similar to the first wave.

## 2 Introduction

On June 29 2020, the World Health Organization documented that more than 10 million cases of infection by SARS-Cov-2 had been reported (World Health Organization, 2020). In continental France, the epidemic seemed to be contained after a series of measures, including a drastic lockdown from March 17 to May 11. During the lockdown, one required a self-authorisation to leave home. The lockdown has successfully slowed down the spread of the epidemic, reducing the reproduction number *R*_*t*_ from around 3 to less than 1 (Salje *et al*., 2020; Sofonea *et al*., 2020; Duchemin *et al*., 2020). The lockdown was then lifted progressively. On May 11, workers were allowed to go back to work, and schoolchildren to go back to school, with new sanitary measures enforced in shops, at work and in schools. Individuals were not allowed to change region or to move more than 100km away from their home. On June 22, all children were expected to attend school, and some of the sanitary measures were no longer enforced. In particular, restrictions on travel within continental France were lifted. As of June 25, the epidemic in continental France is considered under control by Santé Publique France (SPF), the governmental agency in charge of monitoring the epidemic (Santé Publique France, 2020). SPF estimates there are 35 new cases per day per 1 000 000 inhabitants. However, the epidemic has not had the same impact on all regions, and as a result the epidemic still does not have the same intensity in all regions. This can be seen in the predicted proportion of infectious individuals per region on July 1st 2020 (Fig. 1). These predictions are based on a Bayesian model that uses the number of SARS-CoV-2 related deaths through time to monitor the epidemic (Duchemin *et al*., 2020). This model was found to have good accuracy and agreed with other models of the epidemic in France.

**Figure 1:**
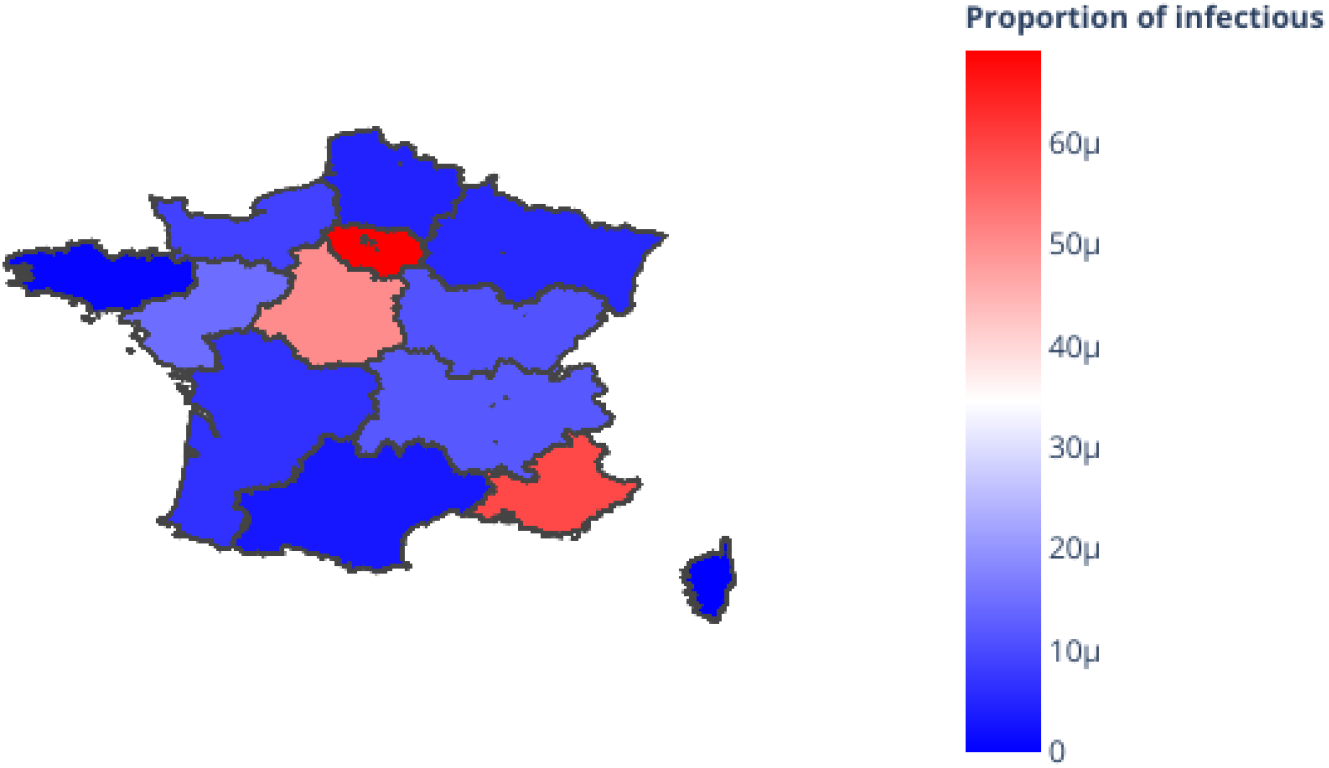
Initial proportion of infectious individuals as of July 1 2020 according to Duchemin et al.’s Bayesian model. One unit *µ* corresponds to one infectious individual per 1 000 000.

SPF also estimates a reproductive number at 0.92 (95% confidence interval: 0.89-0.96) for continental France for the week preceding June 25. Such a low *R*_0_ is due to sanitary measures that are still in place, such as the prohibition of large gatherings or advice on hand washing and mask wearing in shops and at work. But it may also be due to SARS-CoV-2 susceptibility to weather conditions (Neher *et al*., 2020; Kissler *et al*., 2020), *i*.*e*. its seasonality. Other coronaviruses are known to be seasonal, with a higher reproduction number in winter than in summer, but it is still unknown how seasonal SARS-CoV-2 may be. In particular, in their investigations of SARS-CoV-2 seasonality, (Neher *et al*., 2020) used a seasonality value of 0.5 for Northern temperate countries, while (Kissler *et al*., 2020) used a seasonality value of 0.13. Thus it is important to assess how these different estimates affect future trajectories for the epidemic in France.

Between 2014 and 2016, on average 119 million people moved between regions during holidays (Direction Générale des Entreprises, 2018) (Fig. 2). Most of these migrations happen during summer, and could help spread the virus between regions, including moving the virus into regions that comparatively had so far been spared by it.

**Figure 2:**
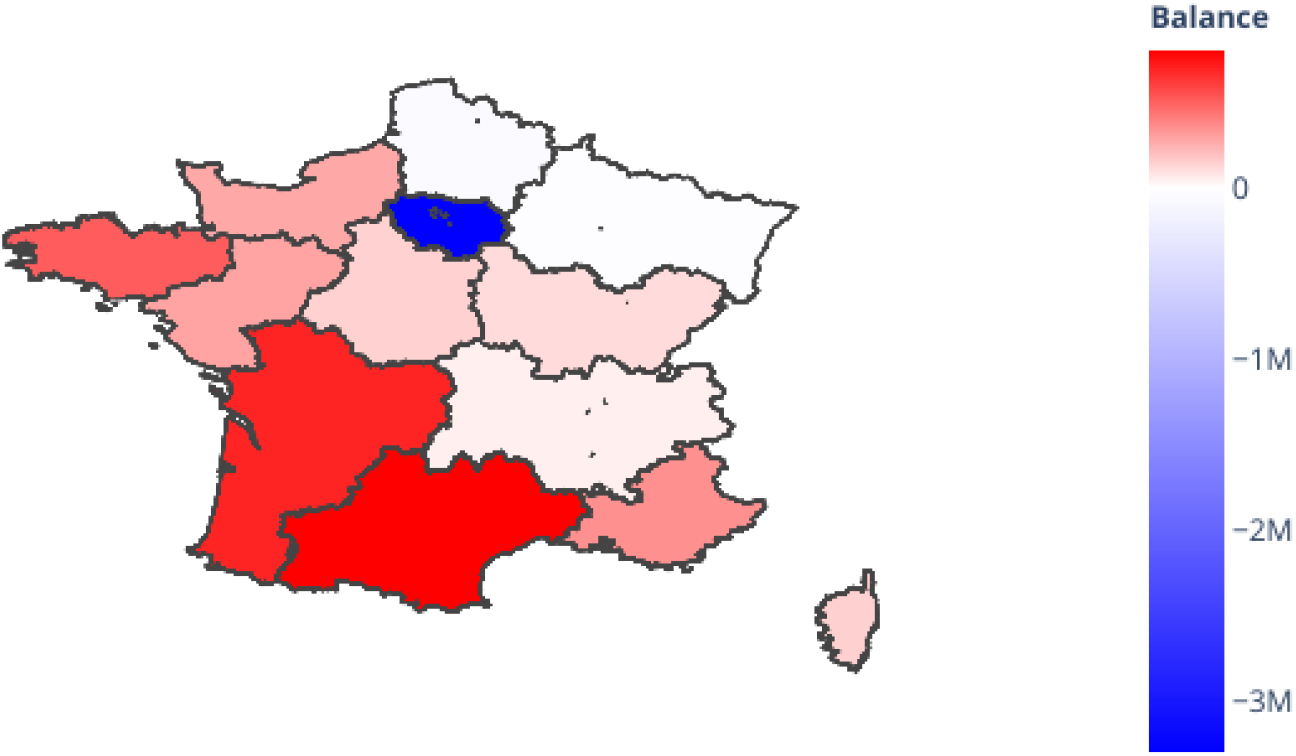
Migration balance during the summer holidays in continental France. The balance between outgoing and incoming migrants is color-coded between blue for regions with negative balance, and red for regions with positive balance.

In this manuscript, we studied the effect summer holiday migrations between regions may have on the spread of the SARS-CoV-2 epidemic in France. We implemented a compartmental Susceptible-Exposed-Infectious-Recovered/Removed (SEIR) model to simulate the infectious process, and complemented it with migrations between regions as gathered from (Direction Générale des Entreprises, 2018). We initialized the number of individuals in each compartment based on simulations from (Duchemin *et al*., 2020)’s model. We first investigated how seasonality and reproduction numbers interact to determine the size and the timing of a second wave of infections in France. To this end, we examined two seasonality values and three reproductive number values for the summer. We finally investigated how migrations affected the number of infections in each region.

## 3 Material and methods

### 3.1 Models

#### 3.1.1 SEIR model

We use a Susceptible-Exposed-Infectious-Recovered/Removed (SEIR) model to simulate the SARS-Cov-2 epidemic in France. Such models are used widely to simulate epidemics, and a formal presentation can be found in (Hethcote, 2000) or (Neher *et al*., 2020). A brief intuitive presentation follows. “Susceptible” individuals may become infected because they coexist with infectious individuals; upon infection, an individual becomes “Exposed”. Exposed individuals then become “Infectious” after an incubation period. Infectious individuals contribute to the spread of the epidemic, and become “Recovered” or “Removed” after an infectious period. Individuals in the R category cannot move to another category.

The starting code base was taken from (Neher *et al*., 2020), who used their SEIR model to investigate the influence of migrations and seasonality on viral spread worldwide. In their model, seasonality and migration parameters were drawn from prior distributions, and other parameters were fixed to values drawn from the literature. They used this model to examine possible trajectories that the worldwide epidemic could take over the next months and years. We built upon this model to investigate the influence of seasonality and summer holiday migrations on the epidemic in France. In particular, we used empirical data to inform the pattern of migrations between French regions in the mainland, and we take into account migrations in both directions: to the holiday region and back. We also used estimated reproduction numbers and combined them with a model of seasonality.

#### 3.1.2 Simulating holiday migrations in the SEIR model

The SEIR model is based on differential equations that describe how individuals move between the S, E, I, R categories in a small amount of time *dt*. Each region has its own set of population size and starting numbers of individuals in the S, E, I, R categories. Other parameters such as the reproduction number or the infectious period are shared between regions. Contrary to agent-based models, each individual in the simulation is not considered separately from other individuals: instead, only the numbers of individuals in each compartment are considered. Simulation is performed by dividing the year in *n* intervals of time *dt* and running the differential equations repeatedly over each interval. Migrations between regions happen every week during the summer holidays. A number of emigrant individuals are sent to destination regions and removed from their origin region; during the move, they keep their S, E, I, R category. These immigrants are added to the numbers of S, E, I, R individuals in the destination region and removed from the numbers of S, E, I, R individuals in the origin region for a week. Then, during a week, these immigrants can change category according to the process happening in the destination region. At the end of the week, the immigrants are brought back to their origin region. The total number of individuals coming back equals the number of individuals that have gone on holidays, but the proportions of S, E, I, R individuals among them can differ from the starting number. With such a process, individuals coming from regions with low infection rates can become infected if they move to regions with high infection rates, and infectious individuals migrating to another region may infect individuals in the destination region.

#### 3.1.3 Joint simulation of seasonality and interventions

To simulate jointly non-pharmaceutical interventions and seasonality, we used estimates of *R*_*t*_ from Duchemin et al.’s Bayesian model and modelled seasonality deterministically. Duchemin et al.’s Bayesian model uses the number of deaths in 13 regions in metropolitan France to estimate an *R*_0_ value before the lockdown, and a set of other parameters that act as modifiers of the initial *R*_0_ to provide estimates of daily *R*_*t*_ during the lockdown, and after the lockdown. These *R*_*t*_ values account for three elements: the effect of seasonality, the effect of other factors, notably non-pharmaceutical interventions, and the decline in the number of susceptible individuals. Taking the average of the *R*_*t*_ values over a time period (*i*.*e*. before, during, or after the lockdown) provides an estimate that we will refer to as *R*_*average*_ here and that factors out the effect of the decline in the number of susceptible individuals. But such an *R*_*average*_ value still combines the effect of seasonality and the effect of non-pharmacetical interventions. The following explains how we disentangle these two factors. We modelled seasonality deterministically as in (Neher *et al*., 2020) as follows:

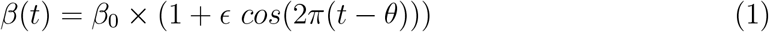

Here *β*(*t*) is the transmissibility at day *t, i*.*e*. the rate at which an infected individual infects another individual, *E* is the strength of seasonal forcing, and *θ* is the date of peak transmissibility. We used the parameter *β*_0_, which was defined as the yearly average transmissibility in (Neher *et al*., 2020) to separate the effects of seasonality and of other factors. To this end, we estimate one *β*_*average*_ value per period, where each *β*_*average*_ value corresponds to an *R*_*average*_ value as follows:

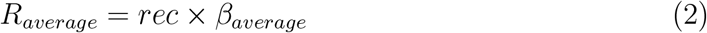

Where *rec* is the recovery rate, *i*.*e*. the inverse of the time for an infected individual to recover. Therefore, to estimate *β*_*average*_ values, we use:

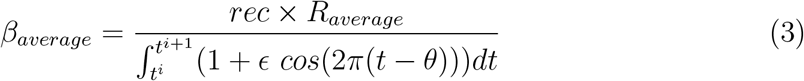

Where *t*^*i*^ and *t*^*i*+1^ are the boundary dates for period *i*. The resulting daily *R*_*t*_ values are shown Fig. 3 for the period between July 1st 2020 and June 1st 2021. These were used as input for the SEIR model, which then handles the decline in the number of susceptible individuals on its own.

**Figure 3:**
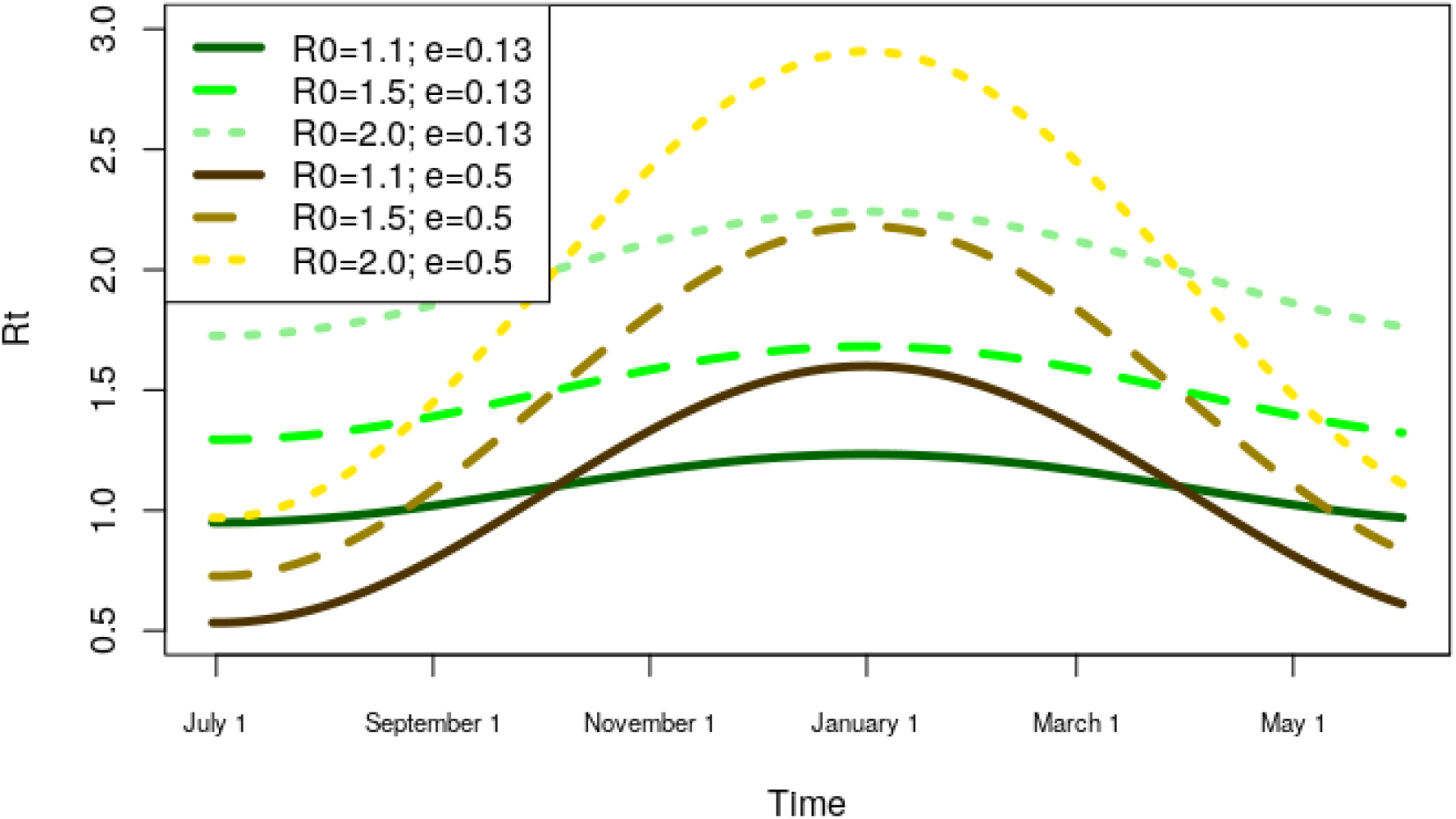
*R*_*t*_ values combining seasonality and different values of *R*_0_. These different trajectories were used as input to the SEIR model.

### 3.2 Data

#### 3.2.1 Parameters

Since (Neher *et al*., 2020)’s work, some parameters of the Covid19 disease have been determined more precisely, but others are still uncertain. In the case of reproduction numbers and seasonality, we compared several values to account for this uncertainty. We acknowledge a certain amount of arbitrariness in selecting the other parameter values but observed little sensitivity of the results when we introduced small variations.

We set the reproduction numbers by combining model-based estimates and seasonality estimates. We use the model-based estimates to provide average reproduction numbers *R*_*average*_ over time-periods, and use seasonality estimates to allow variation around these average values. We explored 3 different *R*_*average*_ values for the period extending from July 1st 2020 to the end of the simulation, June 1st 2021: 1.1, 1.5 and 2.0. We did not explore values below 1.0 as they would be certain to lead to an extinction of the epidemic.

We considered two values for the seasonality of SARS-CoV-2. First we used a value of 0.5, in agreement with (Neher *et al*., 2020)’s analyses of the seasonality of other coronaviruses in Northern temperate countries. Second we used a lower value of 0.13, in agreement with (Kissler *et al*., 2020). In both cases, we considered that the maximum infectivity of the virus was on January 1st.

We set the infectious time, the time it takes between infection and becoming infectious, to 3 days, assuming it takes ≈ 5 days from infection to symptom onset (Lauer *et al*., 2020), and infectiousness starts ≈ 2 days before symptom onset (He *et al*., 2020). This is the time an individual remains in the *E* compartment.

We set the recovery time, the time it takes between symptom onset and the end of infectiousness, to be 6 days, based on the inferred infection profile in (He *et al*., 2020). This is the time an individual remains in the *I* compartment.

#### 3.2.2 Initialization

Region wise proportions of S, E, I, R individuals on July 1 2020 were simulated using a Bayesian model of the French epidemic (Duchemin *et al*., 2020). We chose to use the model without mixture between regions as it provided predictions that were very similar to the mixture model and was easier to fit and simulate. This model was fitted with data up to June 29. The parameter values of each sample of the MCMC chain were used to simulate the epidemic spread up to July 1st 2020. Median values were extracted to initialize the SEIR model in each region. Exposed individuals were defined as individuals that had been infected less than 3 days ago. Infectious individuals were defined as individuals that had been infected less than 9 days ago but more than 3 days ago. Recovered/Removed individuals were defined as individuals that had been infected more than 9 days ago. All the other individuals were assigned to the Susceptible category. This assignment of individuals to categories is consistent with the SEIR parameters introduced above. The proportions of infectious individuals on July 1 2020 estimated by the model range between 3.1 × 10^−8^ for Corsica and 6.9 × 10^−5^ for Île de France, with a mean at 1.9 × 10^−05^. These numbers are in the same range as the number of new cases per day estimated by SPF in the week preceding June 25, which was 3.5 × 10^−5^.

### 3.3 Validation

To validate our implementation of the SEIR model, we ran it between March 1 2020 and June 1 2021 and compared the number of infectious individuals through time to the numbers obtained by running (Duchemin *et al*., 2020)’s Bayesian model over the same time interval. The SEIR model was initialized with the parameter values that the Bayesian model has inferred for March 1 2020.

### 3.4 Implementation and availability

The simulation is implemented in Python and is available at https://gitlab.in2p3.fr/boussau/futurecorona

## 4 Results

### 4.1 Differences between simulated numbers of infectious individuals using the SEIR model and estimates from a Bayesian model

Fig. 4 shows that the SEIR model, when initialized with values drawn from (Duchemin *et al*., 2020)’s model on March 1 2020, qualitatively captures the differences between regions that the Bayesian model infers. However, it dampens the variations that the Bayesian model infers: maxima are not as high in the SEIR model as in the Bayesian model, and minima not as low. These differences between the two approaches are significant and show that the SEIR model lacks in realism. In particular, it would seem adventurous to try and use it to obtain quantitative predictions about the future of the epidemic in France. However, we expect that despite these shortcomings, we can still use the SEIR model to obtain a qualitative picture of what the future of the epidemic may look like.

**Figure 4:**
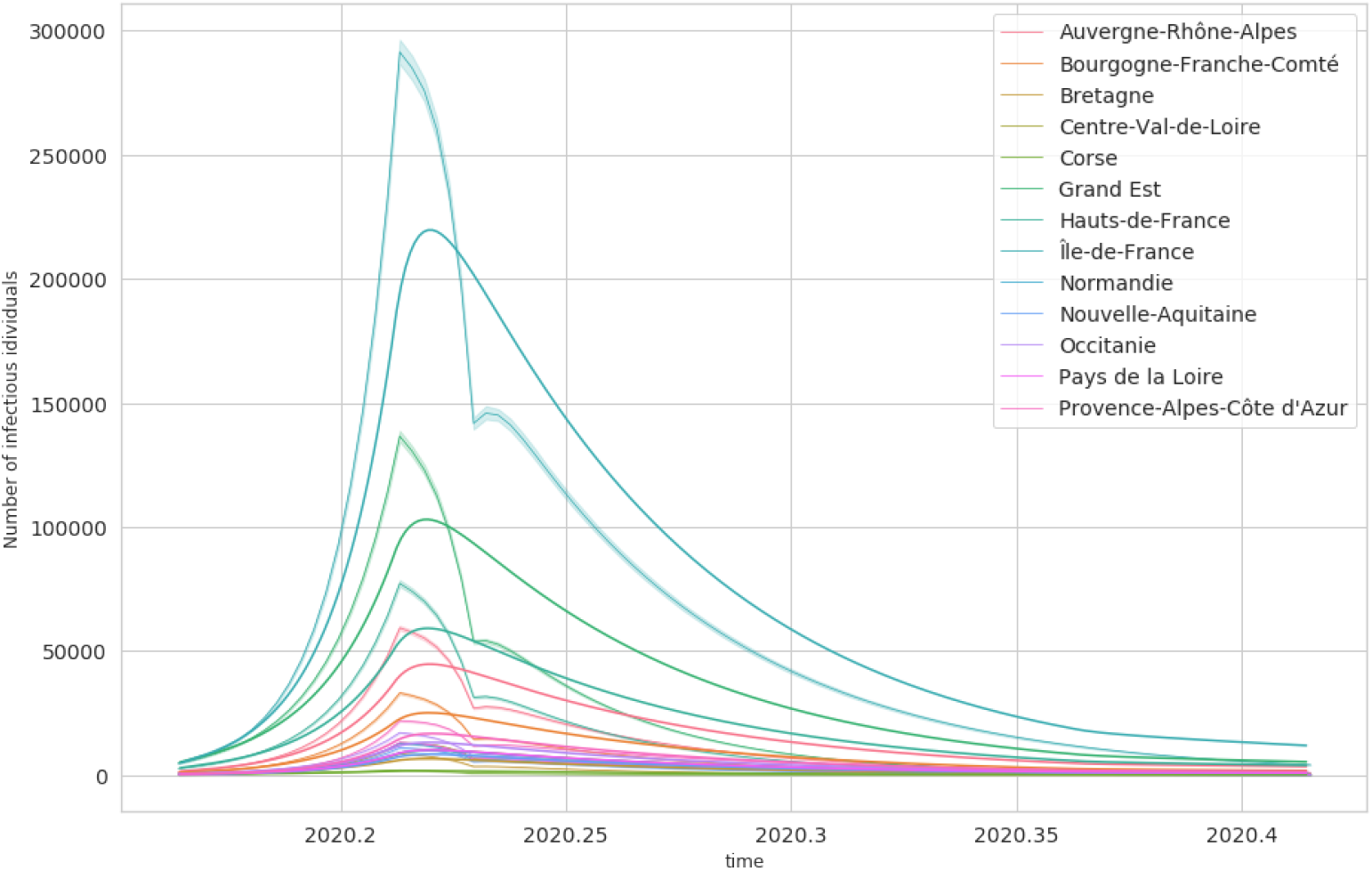
Number of infectious individuals through time according to our SEIR model and according to a Bayesian model. Region-wise estimates from the Bayesian model are represented with a solid line on top of a shaded ribbon, and estimates from the SEIR model are represented with solid lines of matching colors.

### 4.2 Migrations have a minor effect on the number of infections

In the following we examine the total number of infected individuals as of September 1 2020, just after the summer holidays, and at the end of the simulation, June 1 2021. Tables 1 and 2 show that summer migrations have a very limited impact on the total number of infections. This limited impact at the national level is also observed at the regional level (data not shown). The impact seems slightly more important for low seasonality initially 1, but then this effect disappears at the end of the simulation 2. We also observed that migrations contribute to making the wave start slightly earlier (data not shown).

**Table 1:**
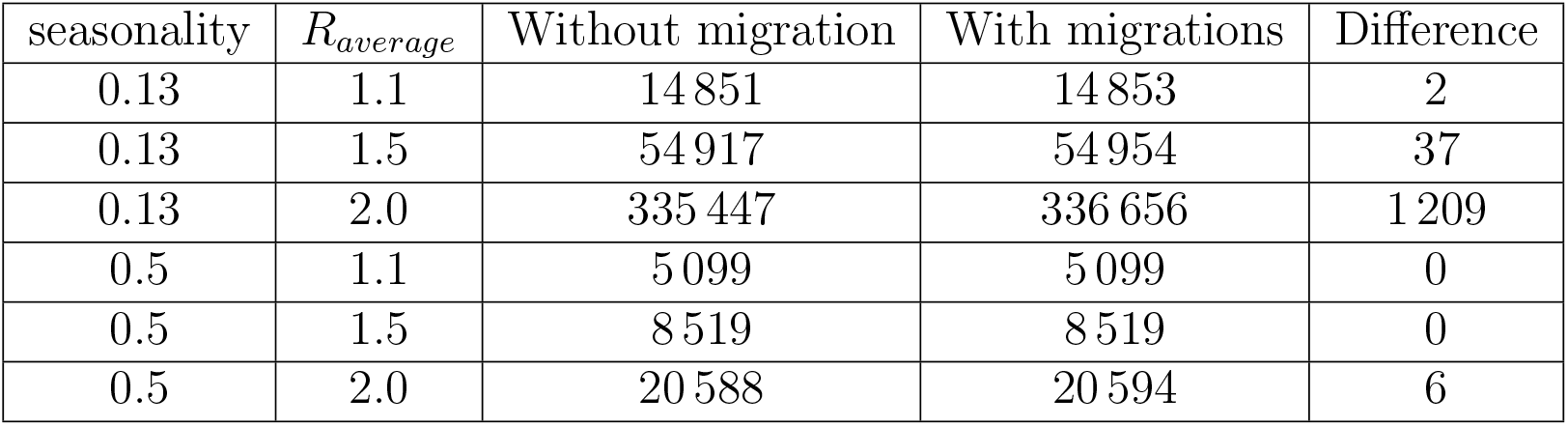
Total numbers of infections as of September 1 2020 for varying seasonality and *R*_*average*_ values.

| seasonality | $R_{average}$ | Without migration | With migrations | Difference |
| --- | --- | --- | --- | --- |
| 0.13 | 1.1 | 14 851 | 14 853 | 2 |
| 0.13 | 1.5 | 54 917 | 54 954 | 37 |
| 0.13 | 2.0 | 335 447 | 336 656 | 1 209 |
| 0.5 | 1.1 | 5 099 | 5 099 | 0 |
| 0.5 | 1.5 | 8 519 | 8 519 | 0 |
| 0.5 | 2.0 | 20 588 | 20 594 | 6 |

**Table 2:** Total numbers of infections as of June 1 2021 for varying seasonality and *R*_*average*_ values.

| seasonality | $R_{average}$ | Without migration | With migrations | Difference |
| --- | --- | --- | --- | --- |
| 0.13 | 1.1 | 994 670 | 995 048 | 378 |
| 0.13 | 1.5 | 42 854 732 | 43 010 354 | 155 622 |
| 0.13 | 2.0 | 53 648 401 | 53 936 387 | 287 986 |
| 0.5 | 1.1 | 1 338 694 | 1 338 879 | 185 |
| 0.5 | 1.5 | 51 795 178 | 51 839 875 | 44 697 |
| 0.5 | 2.0 | 59 889 135 | 60 208 067 | 318 931 |

### 4.3 A second wave is observed in all parameter settings

The possibility of a second wave of infections depends on the strength of seasonality for SARS-Cov-2, and on its reproduction number *R*_0_. Fig. 5 shows the number of infectious individuals through time according to 2 different values for seasonality (low: 0.13, high: 0.5), and 3 different values for *R*_*average*_ (1.1, 1.5, 2.0). Higher *R*_*average*_ values result in earlier and steeper waves, because the epidemic spreads fast. When *R*_*average*_ = 1.1, the wave is limited, peaking at 229 000 infectious individuals for low seasonality. Such a wave would be comparable to the first wave, which peaked at 307 000 individuals on March 29 according to (Duchemin *et al*., 2020)’s model fitted on data up to June 29. Higher seasonality results in later and steeper waves. This is because, in our simulation, for a given *R*_*average*_, *R*_0_ values of high seasonality scenarios are inferior to the *R*_0_ values of low seasonality scenarios (Fig; 3). At the earliest, for low seasonality and high *R*_*average*_, the second wave peaks in October 2020; at the latest, for high seasonality and medium *R*_*average*_, the second wave peaks in January 2021 or even April 2021 for low *Raverage*.

**Figure 5:**
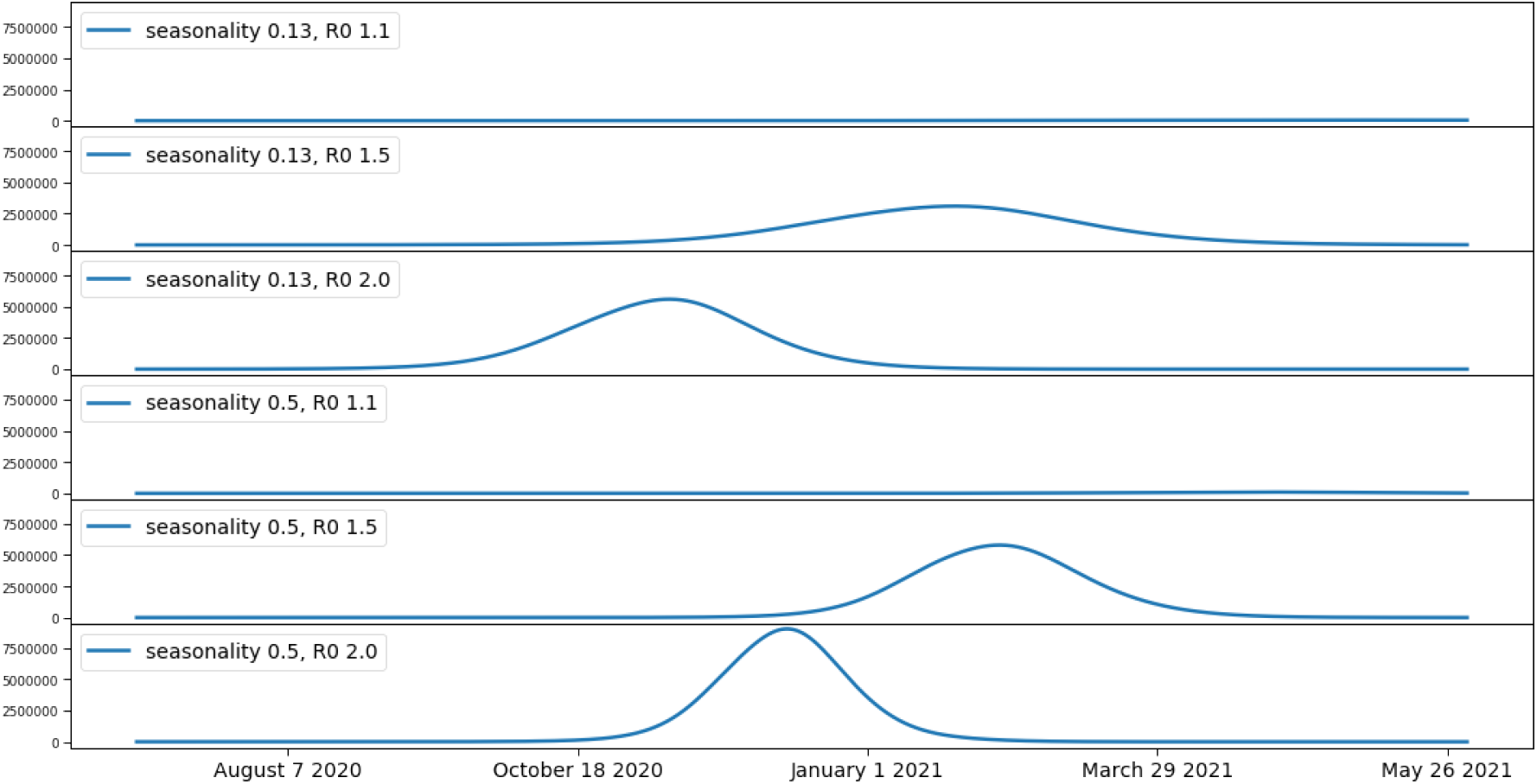
Number of infectious individuals through time. Scenarios with and without migration provide indistinguishable curves. 3 top lines: weak seasonality, *R*_0_ at 1.1, 1.5, 2.0 from the top to the bottom. 3 bottom lines: strong seasonality, *R*_0_ at 1.1, 1.5, 2.0 from the top to the bottom.

## 5 Discussion

In this report we investigated the effect of several parameters on a model of the SARS-CoV-2 epidemic in France. We varied the value of the reproduction number, the value of the seasonality, and took into account summer holiday migrations in a SEIR model. Our choices of 1.1, 1.5, 2.0 for the future reproduction number of SARS-CoV-2 correspond to three hypotheses. In all cases, they assume that the sanitary measures currently in place, which include mandatory mask wearing in public transportation and at work, and the prohibition of large gatherings, are not sufficient to keep the reproduction number below 1.0, which means that the epidemic can spread. All choices are lower than the estimated value of the *R*_*average*_ before the lockdown was enforced, which is around 3.0 (Duchemin *et al*., 2020; Salje *et al*., 2020; Sofonea *et al*., 2020), which conveys the idea that the sanitary measures do reduce the *R*_*average*_. The other parameters of the model were either obtained from estimates by (Duchemin *et al*., 2020)’s Bayesian model, or drawn from the literature. Variations in these parameters can affect the timing and the scale of the second epidemic wave (not shown), but unless extreme values are used, do not make it disappear.

We chose to use a deterministic SEIR model to quickly explore a range of scenarios. We do not attempt to take into account the uncertainty in most parameters, and do not attempt to model the inherent stochasticity of the epidemic process. The SEIR model assumes that all individuals behave the same way and have the same characteristics.

In particular, it assumes that all individuals in a given compartment and region have the same probability to be infected or to infect another individual. Models that allow heterogeneity between individuals have been proposed and can alter the predicted dynamics of the epidemic (Gomes *et al*., 2020). It is possible that such models may fit the dynamic of the epidemic better than our model, whose shortcomings are apparent Fig. 4. Future work may address the effect of migrations with such models. In the mean time we expect that our SEIR model can capture important aspects of the epidemic. The predictions of our model may provide insight into possible central trajectories of the epidemic, but cannot be used to infer e.g. a detailed timing of the progress of the epidemic.

Although the different French regions had different proportions of infectious individuals on July 1 2020 (Fig. 1), and the numbers of migrating individuals were large, we observed that holiday migrations have little effect on the epidemic (Tables 2, 1). Our results are conservative, because we used an overestimate of the number of migrants during the summer holidays, which corresponds to the average number of migrants per year over 2014-2016, and not just during summer holidays. During migrations, at most 29 783 infectious individuals changed region on a given week (for *R*_*average*_ = 2.0 and low seasonality). Compared to the 64 million individuals in France, these inputs were not sizeable enough to alter the dynamic of the epidemic in a meaningful way. This reasoning also explains why low seasonality results in a slightly larger impact of holiday migrations: in this condition higher summer *R*_0_ values allow the epidemic to start an epidemic wave, meaning that the number of infectious migrants become less and less insignificant as time goes by. This shows that it is important that the number of infectious migrants remains low for migrations to have a limited impact on total numbers of deaths. However, small numbers of infectious migrants could still contribute to creating local clusters of infections. Investigating the likelihood of such clusters would require using an agent-based model with a realistic and detailed map of where individuals spend their holidays.

In a recent manuscript, (Britton & Ball, 2020) investigated the effect of summer migrations from a densely populated metropolitan area, Stockholm, to a popular summer vacation destination, the island of Gotland. They used a SIR model in which they introduced two types of visitors, long term visitors and short term visitors, assuming short term visitors have more contacts with the locals. They investigated a range of parameter values and found that migrations can increase the number of infections on the island. However, they also found that this increase depends upon the number of infectious migrants. Their results support our conclusion that low numbers of infectious migrants are key for migrations to have a limited impact on total numbers of deaths.

The reproduction number and the seasonality parameter have a large effect on the epidemic, and control the size, the spread and the date of a second wave. Even if the reproduction number is low at 1.1, a second wave is obtained. If the reproduction number is larger, the second wave happens earlier and is steeper and larger. This second wave is unsurprising given the low proportion of immunized individuals after the first wave, and given the historical precedent of the 1918 Influenza (Taubenberger & Morens, 2006).

## 6 Conclusion

We investigated the influence of summer holiday migrations and of various parametrizations for the seasonality and the reproduction number on the epidemic of SARS-CoV-2 in France. To this end, we modified a SEIR model to allow for migrations between regions during the summer holidays. We found that migrations of French individuals during summer holidays have almost no impact on the epidemic. This is a consequence of the lockdown which has been successful in decreasing the number of infectious migrants. Different seasonality and reproduction numbers above 1.0 all result in a second wave, whose peak date can vary between October 2020 and April 2021. Such a second wave would be avoided if the sanitary measures currently in place in France are sufficient to keep the reproduction number below 1.0. If the sanitary measures manage to keep the reproduction number low, for instance at 1.1 as in one of our simulations, the second wave is flattened and could be similar to the first wave. Such a flatter wave would be much more manageable for the French health system, hence it important to keep the reproduction number low.

## Data Availability

The data and scripts used in this manuscript are available on a public repository.

https://gitlab.in2p3.fr/boussau/futurecorona

## 7 Author contributions

LD provided code for running the Bayesian model to initialize the simulations. MP contributed to data analysis and acquisition. All authors discussed the ideas in the manuscript, and read and commented on the manuscript. BB directed the work, implemented the model and wrote the manuscript.

## 8 Acknowledgements

The authors would like to thank (Neher *et al*., 2020) for implementing their model in open source, which sped up our research, and Philippe Veber for fruitful discussions.

